# T cell transcriptional and receptor signatures predict response to telomerase vaccination in prostate cancer

**DOI:** 10.64898/2026.05.25.26354038

**Authors:** Eirik Høye, Reetta Nätkin, Karishma Sajnani, Nikolai Engedal, Julia Elisabeth Simensen, Sini Hakkola, Antti Kiviaho, Francesco Ballesio, Thomas Cecchetto, Espen Basmo Ellingsen, Marita Westhrin, Eivind Hovig, Anthony Mathelier, Tapio Visakorpi, Teuvo L. Tammela, Teemu Johannes Murtola, Sini Eerola, Matti Nykter, Wolfgang Lilleby, Alfonso Urbanucci

## Abstract

While prostate cancer (PC) is defined as immunologically cold, limiting the efficacy of immune checkpoint inhibitors, therapeutic vaccination targeting tumor-associated antigens represents an attractive strategy to promote disease control in low volume metastatic patients. The UV1 cancer vaccine is based on immunization with tripeptide fragments from human telomerase reverse transcriptase (hTERT) and a phase II clinical trial demonstrated induction of robust T cell response in men with *de novo* metastatic castration-sensitive prostate cancer (mCSPC). Comparison with long-term survival data of non-metastatic CSPC patients as reference showed that despite metastatic disease at diagnosis, UV1-treated patients who mounted an early vaccine-induced immune response achieved progression-free and overall survival comparable to non-metastatic patients. We examined biological determinants of clinical benefit following UV1 vaccination including tumor transcriptome and T cell receptor (TCR) profiling from circulating and tissue resident T-cells of the 22 men enrolled. Analysis of diagnostic and post-UV1 treatment biopsies revealed that low baseline exhaustion of T cells and higher CD8^+^ T cell abundance are associated with early immune response to the vaccine and longer survival. Moreover, we identified specific TCR motifs relative to early responders, that can indicate potential benefit from UV1 vaccination.

These findings indicate that baseline intratumoral T cell exhaustion state and repertoire shape responsiveness to hTERT vaccination and long-term outcome. Overall, our study underlines how baseline immune profiling may be used as a companion biomarker to predict mCSPC patients most likely to benefit from therapeutic vaccination.

## Introduction

For *de novo* metastatic castration-sensitive prostate cancer (mCSPC), standard initial management now involves intensified systemic therapy combining androgen deprivation therapy (ADT) with other treatments including androgen receptor pathway inhibitors, chemotherapy and incorporating radiotherapy (RT) against the primary tumor (1). The clinical trial STAMPEDE showed that irradiation to the prostate results in clinical benefits in low volume metastatic diseases (2). The underlying mechanism of this benefit is not well understood; however, the data suggests that RT can enhance anti-tumor immunity by releasing damage-associated antigens which can activate innate and adaptive immune response mechanisms (3).

In this setting, cancer vaccines which leverage mRNA, peptides and dendritic cells are being evaluated as adjuvant treatments with increasing evidence of showing the potential to overcome tumor–mediated immunosuppression with antigen-specific T cell responses that can synergize with chemotherapy, RT, or checkpoint inhibitions (4). In prostate cancer, sipuleucel-T remains the only FDA approved immunotherapeutic cancer vaccine for metastatic castration-resistant patients and has demonstrated limited survival benefit (5), although novel peptide-and DNA-based vaccines targeting prostate antigens are being evaluated (6).

We have previously demonstrated that hTERT-derived peptides are potent activators of CD4⁺ T helper (Th) cells, and that vaccination with the UV1 hTERT peptide vaccine, hereby named UV1, elicits Th immune responses in a high proportion of oligometastatic *de novo* metastatic (M1) castration-sensitive prostate cancer patients (NCT01784913) treated with ADT and RT (7), with vaccine-specific immune responses frequently induced early after treatment initiation and, in some patients, sustained for several years without evidence of clinical relapse patients (8,9). The molecular mechanisms underlying such durable treatment responses, particularly in patients who experienced immune response within 10 weeks of the first vaccination, remain elusive.

We recently demonstrated the potential to stratify (PC) patients for treatment response using whole transcriptome profiling (10). The study did not include detailed immunoprofiling, which may be critical for identifying patients most likely to benefit from immunotherapy including cancer vaccines. In fact, T cell receptor (TCR) repertoire analysis may also provide insight into patient stratification (11), although the biological mechanism underlying activation of adaptive immunity in PC remains elusive (12–14).

In this study, we integrated transcriptome profiling and TCR repertoire analysis of diagnostic and post UV1 treatment tumor biopsies to investigate tumor and immune features associated with response to UV1 vaccination in mCSPC patients. By comparing *de novo* metastatic patients treated with UV1 to a non-metastatic reference cohort, we characterized immune determinants of vaccine responsiveness. We find that lower T cell exhaustion and higher abundance of a set of TCR sequence motifs in tissue resident T cells at diagnosis are associated with early immune responses to the vaccine and may explain the durable remission in these patients at more than 10 years follow up.

Our findings suggest that a more potent activation of vaccine-specific T cells in early immune responders is due to pre-existing immune features in these patients. Moreover, we demonstrate that deep immunoprofiling of metastatic CSPC patients, together with analysis of their tumor-infiltrating T cell receptor repertoires, can enable development of predictive classifiers for patients benefiting from similar therapeutic cancer vaccine approaches or immunotherapy.

## Methods

### Clinical Material

The UV1 phase I/II trial (NCT01784913) investigated hTERT peptides vaccine in men with androgen-sensitive metastatic PC (*n* = 22). The institutional protocol was approved by the Ethical Committee (EC) Health South–East Region (EudraCT 2012-002411-26). Patients referred to the study started ADT (luteinizing hormone-releasing hormone agonist, Zoladex^®^ 10.8 mg subcutaneously every third month) and received bicalutamide 50 mg per day at enrolment until progression. Patients were recruited independent of their HLA type. The primary tumor was treated at the local Oslo University Hospital (OUH) with definitive RT. Phenotypic and functional characterizations on patient-derived vaccine-specific immune responses is described in (7) (See **Supplementary Figure 1**). We obtained FFPE embedded diagnostic biopsies from all the men enrolled in the UV1 trial and from 22 men with non-metastatic CSPC enrolled in intensity modulated RT (IMRT) trial. Details for the IMRT trial are provided in **Supplementary Methods**. We also obtained biopsies specimens post UV1 vaccination commencement from the OUH biobank and originally collected by Dr Lilleby at the time of intraprostatic fiducial insertion (start of RT treatment) according to (7). Blood samples from patients enrolled in the UV1 trial were collected prior to beginning of vaccinations and after beginning of vaccinations, on weeks 1, 2, 6, 10, 14, 18, 22, and 26 and every 6 months thereafter as described in (9) (See also **Supplementary Figure 1**).

### DNA and RNA extraction and sequencing

DNA and RNA were simultaneously extracted from paraffin-embedded needle biopsy specimens using the truXTRAC FFPE total NA Plus Kit - Column (Covaris, CAT#PN520252) and following the kit protocol with slight modifications (See **Supplementary Methods**). DNA was used to sequence TCRs from prostate tissue at Cegat GMbH genetic diagnostics and NGS services, targeting the beta chain rearrangements of T cell receptors, including CDR3 region subsequently sequenced on the Illumina platform (Read length: 2 x 100 bp, Output: 2 M clusters (0,4 Gb) per sample). TCR sequencing from peripheral blood is described in **Supplementary Methods**. For transcriptome analysis, RNA was subjected to 3’ end QuantaSeq Library preparation and sequencing with MySeq at the genomic NorSeq core facility at OUH.

### Transcriptomic and immune signature analyses

RNA sequencing analysis was performed on pretreatment tumor biopsies from patients treated with the UV1 while paired post-treatment biopsies were available for 13 patients. RNA-seq data were processed using a standardized pipeline including quality control, trimming, alignment to hg38, and gene-level quantification (see **Supplementary Methods** for full details). Gene signature activities were calculated from quantile-normalized expression data using curated gene sets representing biological pathways, transcription factor targets, and T cell exhaustion programs (**Supplementary Table 1**). Gene set variation analysis (GSVA) was applied to estimate per-sample signature enrichment scores. Associations between pretreatment signature activities and progression-free survival (PFS) or prostate cancer–specific mortality (PCSM) were evaluated using Cox proportional hazards models and Kaplan–Meier analyses. Immune cell-type fractions were inferred from bulk transcriptomic data using EPIC (15)and analyzed associations with survival outcomes. Full statistical procedures are described in the Supplementary Methods.

### TCR repertoire analysis and clonal expansion

T cell clonotypes were clustered into sequence motifs using ClusTCR (16) based on CDR3 similarity, generating a unified network across the full cohort (see **Supplementary Methods** for detailed parameters). TCR diversity was assessed using Hill diversity profiles (17). Differentially abundant TCR motifs between early, late, and non-responders were identified using DESeq2(18). Motifs significantly enriched in early responders (FDR < 0.05) were defined as early-responder signature motifs. Clonal overlap between samples was quantified using the Morisita–Horn Index (19).

Longitudinal PBMC samples were analyzed to assess clonal expansion and persistence. Expanded clones were defined based on log2-fold change and persistence across follow-up samples. Sequence convergence analyses were performed using network-based metrics derived from Levenshtein distance comparisons (20). Full computational and statistical details are provided in the **Supplementary Methods**.

### Spatial VDJ and sequencing

The protocol is described in more details in **Supplementary Methods**. Briefly, cDNA before any fragmentation step was subjected to 16 hours of hybridization-capture–based enrichment targeting CDR3 region of immunoglobulin (IG) and T cell receptor (TR) constant regions following Engblom et al. (2023) (21). We utilized full length spatial barcoded cDNA also profiled with spatial transcriptomics processed on 10x Genomics Visium 3′ arrays to generate spatially barcoded cDNA, as described in Kiviaho et al. (2024) (22). The cDNA was derived from four sections of clinical specimens, representing one benign prostatic hyperplasia (BPH), one treatment-naïve (TRNA) primary PC, one ADT neoadjuvant-treated (NEADT) primary PC, and one locally recurrent castration-resistant prostate cancer (CRPC). After hybridization-capture–based enrichment the cDNA was subjected to library preparation with SMRTBell kit 3.0 from PacBio and samples were sequenced on Revio SMRT cell for PacBio long read sequencing at the the DNA Sequencing and Genomics Laboratory (BIDGEN) in Helsinki. Demultiplexed PacBio reads were processed using a custom analysis pipeline implemented in Python. Reads were filtered based on the presence and orientation of expected library adapter and template-switching sequences, which were used to define read boundaries and standardize read direction (Full details are in the **Supplementary Methods**).

## Results

### Early UV1 vaccine response aligns survival of *de novo* metastatic patients with mon-metastatic disease and reveals shared tumor transcriptomic features

The UV1 trial was a single-arm trial combining standard RT and ADT with UV1 vaccination in men with mCSPC using a three peptides fragments of hTERT (23). Since a vaccine-induced immune response within 10 weeks from first vaccination (early response) was associated to significant better outcome than in patients experiencing late or no immune response (7), we first sought to assess whether early immune responders obtained meaningful benefit from the vaccination. To do so, we compared long-term outcomes between the UV1 metastatic cohort and the IMRT non-metastatic cohort since the IMRT trial run in the same hospital, and since in both trials comparable RT and ADT regimens were used, and differed only in metastatic status (M1 vs M0 disease) of the patients at diagnosis, and the addition of the UV1 vaccination.

At 10-year follow up, timing of the immune response clearly modified the survival probability. For metastatic patients who mounted an early immune response to the UV1 vaccine, despite having metastatic disease, median survival was similar to the IMRT M0 cohort (not reached in 150 months vs median 170 months, respectively; **Figure 1A-B**). The early immune responders’ group derived significant benefit from treatment, with survival probability comparable to those of non-metastatic patients in the IMRT cohort (0.625 CI 0.365-1.000 (PCSM) vs 0.824 CI 0.749-0.907 (PCSM) for IMRT and p = 0.9) (**Figure 1A-B** and **Supplementary Figure 2A-B**). These findings suggest that either the vaccination induced biological effects, or intrinsic patient characteristics contributed to the observed benefit. In contrast, late immune responders had a median survival of 49 months only (0.0917 CI 0.0142-0.5931 (PCSM) vs 0.824 CI 0.749-0.907 (PCSM) for IMRT and p = 1 x 10^-11^), consistent with reported outcome for patients with metastatic disease at diagnosis (24,25).

**Figure 1.**
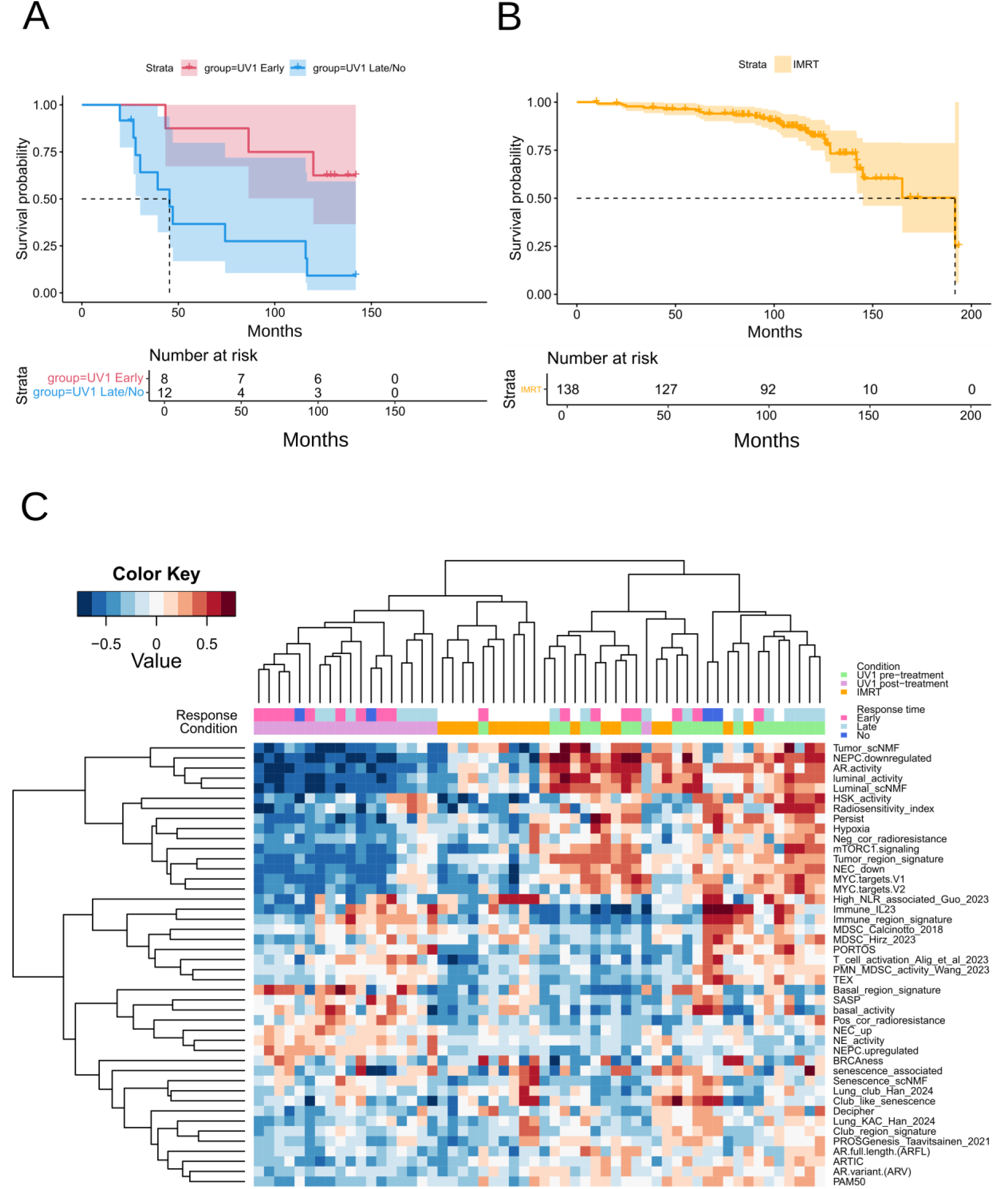
Clinical outcomes of UV1-treated patients compared with an IMRT cohort. (**A**) Kaplan-Meier analysis of prostate cancer-specific mortality (PCSM) comparing metastatic castration-sensitive prostate cancer patients vaccinated with the UV1 tripeptide vaccine (UV1 cohort) and that experience early immune response (<10 weeks from first vaccination), and late or non-responders (> 10 weeks from first vaccination; p = 0.007) (**B**) Kaplan-Meier analysis of PCSM in non-metastatic castration sensitive prostate cancer patients treated with intensity-modulated radiotherapy (IMRT cohort). (**C**) Unsupervised clustering of gene set variation analysis (GSVA) based activity scores of the indicated molecular classifiers (gene signatures; See Supplementary Table 1) across patients in IMRT and UV1 cohorts’ biopsies. For the UV1 cohort, diagnostic biopsies are labelled as “UV1 pre-treatment” while a post UV1 vaccination regime biopsy is labelled as “UV1 post-treatment”. Immune response (early, late, or no response) to the vaccine is also indicated.

To examine whether intrinsic tumor biology features of the UV1-treated patients led to benefit from the vaccination, we profiled the tumor transcriptome using the patients’ core biopsies from both IMRT and UV1 trial cohorts. Principal component analysis (PCA) of the diagnostic biopsies did not clearly separate early and late responders; however, we observed a clear separation of samples pre-versus post-UV1 vaccination (**Supplementary Figure 2C**). Furthermore, diagnostic biopsies in the UV1 (M1) and IMRT (M0) cohorts revealed that the transcriptomic profiles were remarkably similar (**Supplementary Figure 2D**).

Next, to study the tumor biology of these patients, we calculated GSVA activity scores (See methods) for all specimens (**Figure 1C**) using a compendium of previously reported transcriptome-based classifiers (10) and assembled gene signatures (**Supplementary Table 1**). IMRT and UV1 diagnostic biopsies clustered together, while post-UV1 vaccination biopsies formed a distinct cluster (**Figure 1C**). A cluster of classifiers depicting androgen receptor (AR) activity, luminal tumor differentiation (e.g. *AR.activity*, *Tumor_scNMF*, *NEPC_downregulated*, *NEC_down*), and proliferation (e.g. *MYC targets* and *Persist*) had higher score in diagnostic biopsies from the UV1-cohort followed by the IMRT cohort and the post-UV1 treatment biopsies, reflecting both biological differences and the effect of ADT. Corroborating these observations, the classifiers *Persist* and *Decipher*, both prognostic in metastatic disease (10), had significantly higher scores in UV1 diagnostic biopsies compared with post-UV1 biopsies (p = 1 × 10⁻⁶ and p = 9 × 10⁻⁴, respectively). Furthermore, *Decipher* scores were significantly higher in late responders than in early responders to UV1 vaccination (p = 0.03) although, overall, the classifiers-based clustering analysis did not clearly separate biopsies of patients experiencing early versus late or no immune response to the vaccination. Remarkably, immune-related classifiers related to immune infiltration and inflammation (e.g. *Immune_region_signature*, *Immune_IL23*), T cell exhaustion (*TEX*), or depicting infiltration of myeloid immunosuppressive cells (e.g. *MDSC_Calcinotto2018* and *High_NLR_associated_Guo*), defined a specific subcluster of classifiers (**Figure 1C**) which elicited increased variability in UV1 patients (both in diagnostic biopsies pre-UV1 and post-UV1 vaccination), suggesting that despite the tumor transcriptome being remarkably similar, elicited higher degree of tumor infiltration in metastatic patients.

Taken together, these findings indicate that mCSPC might benefit from UV1 vaccination and transcriptome classifiers analysis from diagnostic biopsies could be used to infer known determinants of tumor biology.

### Transcriptomic analysis of biopsy samples identifies T-cell exhaustion as a key feature of the immune response to the UV1 cancer vaccine in metastatic patients with prolonged survival

To characterize clusters of relevant gene sets discriminating patients’ outcomes, we first correlated all the GSVA scores separately in the two patient cohorts (**Figure 2** and **Supplementary Figure 3**). Similar high correlations were independently observed among the signatures in the two cohorts; however, the correlations were stronger in the UV1 cohort than in the IMRT cohort (**Supplementary Figure 3**). Luminal cell-type signatures, proliferation, and MYC-related signatures were highly correlated within the UV1 cohort, and these also strongly correlated with AR activity (**Figure 2**) while in the IMRT cohort the correlations between differentiation markers and AR activity were weaker. Basal and club cell signatures also showed strong correlations, as did inflammatory infiltration and T cell exhaustion signatures (**Figure 2**), but these were weaker in the IMRT cohort (**Supplementary Figure 3**).

**Figure 2.**
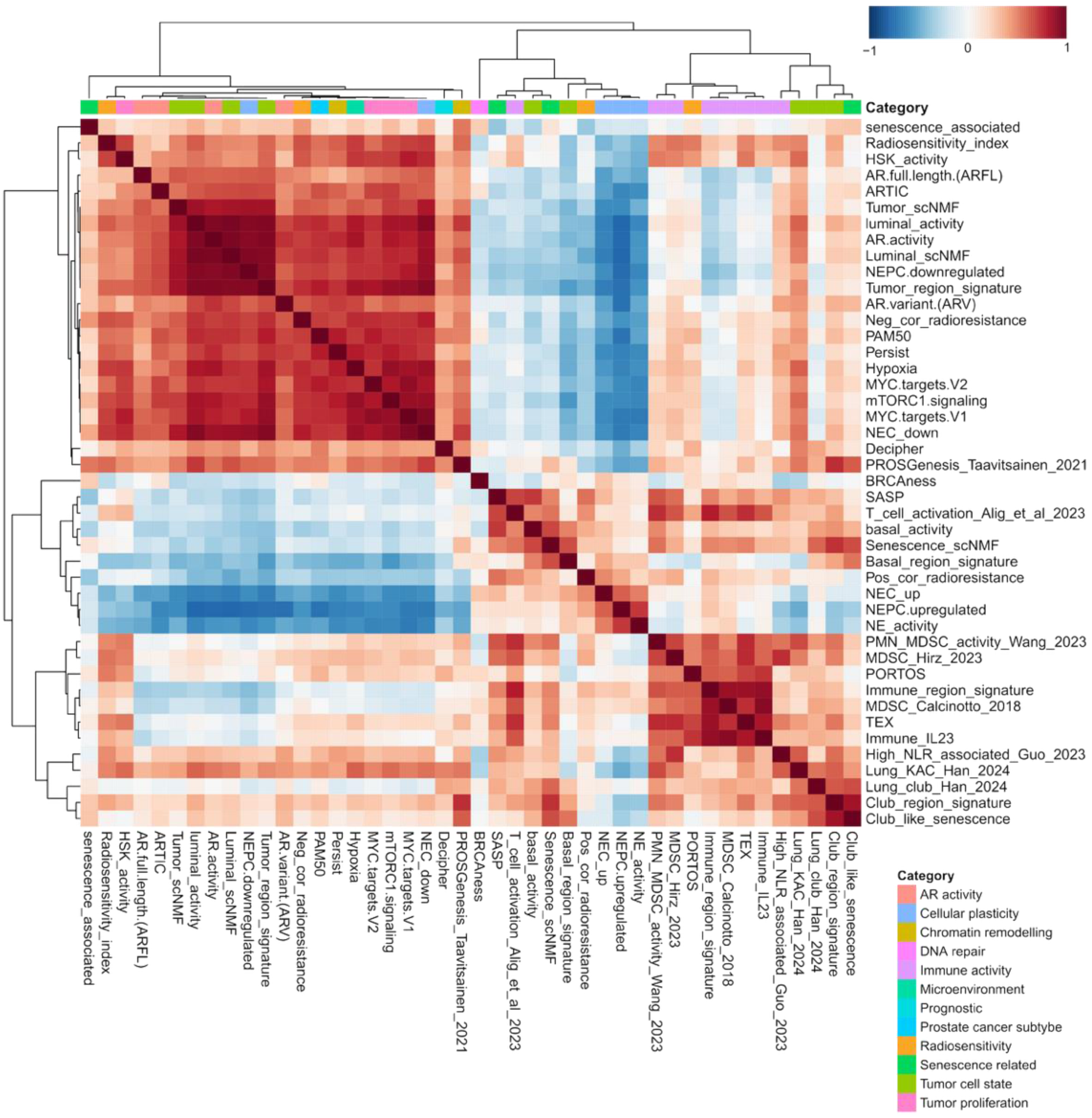
Transcriptome classifiers performance in the UV1 cohort. Unsupervised clustering of gene set variation analysis (GSVA) signature scores (see Supplementary Table 1) based on Pearson correlation in diagnostic biopsy specimens obtained prior to vaccination in the UV1 cohort. Associations between selected signature scores and prostate cancer–specific mortality (PCSM) are shown for the UV1 cohort.

These results indicate that signature activity scores reflect coordinated tumor biology driving aggressive phenotypes. This observation is consistent with recent studies demonstrating that such processes are tumor-intrinsic and established prior to therapeutic intervention (10).

We then assessed the relationship between transcriptome classifiers scores and prostate cancer specific mortality (PCSM) and progression-free survival (PFS), in both the UV1 (**Figure 3A-B** and **Supplementary figure 4A**) and IMRT cohorts (**Figure 3B** and **Supplementary figure 4B**). The T cells exhaustion (TEX) (26) signature score in the UV1 cohort showed a strong positive correlation with poor clinical outcomes, including PCSM and PFS, whereas AR activity was significantly positively correlated with outcomes. These associations were specific to the UV1 cohort and were not observed in the IMRT cohort.

**Figure 3.**
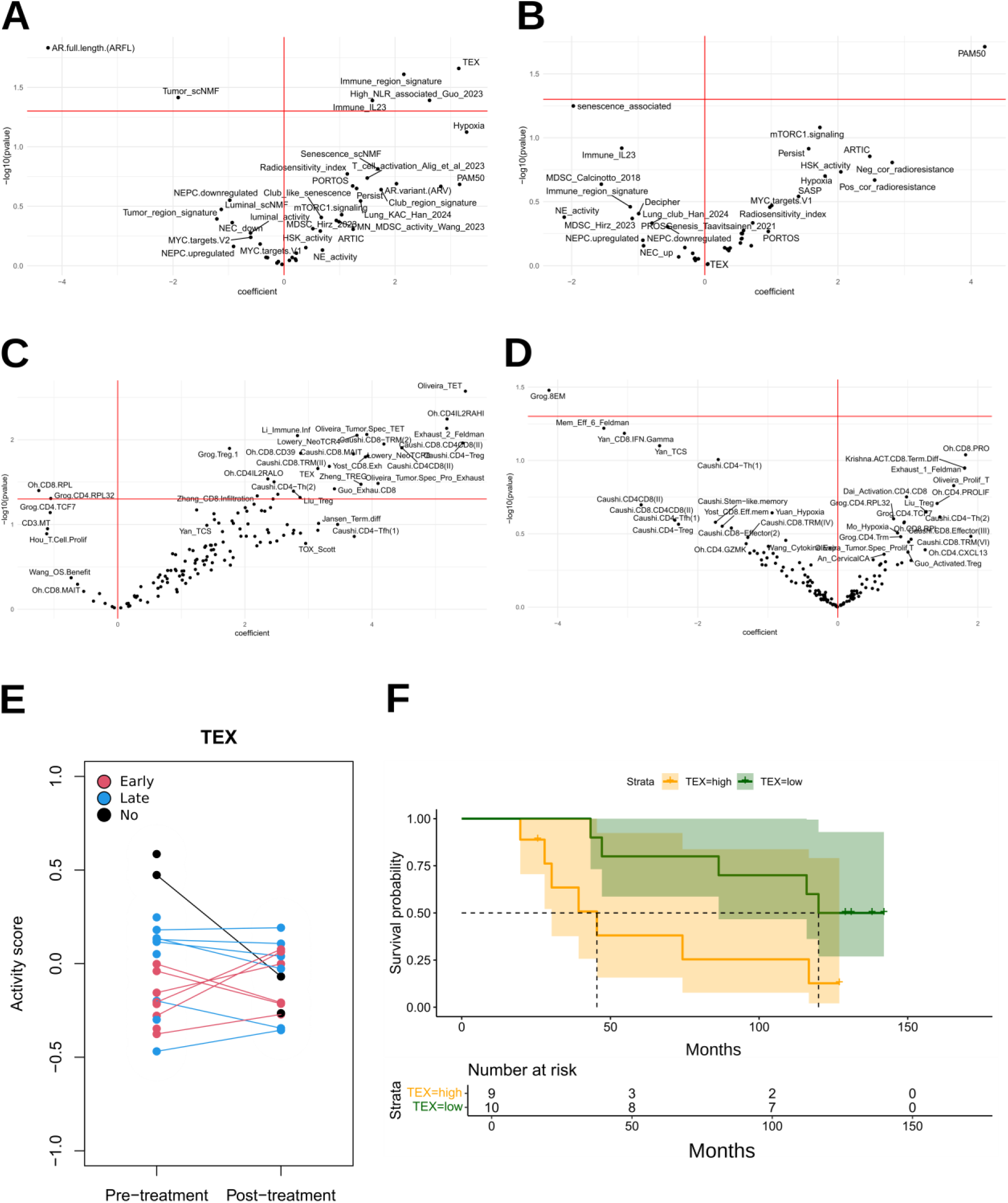
T cell exhaustion is associated with prostate cancer–specific mortality and immune response to cancer vaccine. Forest plots showing the association between gene signature activity and prostate cancer–specific mortality (PCSM) in the UV1 cohort **(A)** and the IMRT cohort **(B)**, with hazard ratios (HRs) displayed on the x-axis. Associations between T-cell exhaustion–related signatures described by Hitscherich et al. (2025) (15) and PCSM are shown for the UV1 cohort **(C)** and the IMRT cohort **(D)**. **(E)** A ladder plot illustrates T cell Exhaustion (TEX) signature activity score (Riegle et al. (28)) in pre UV1 treatment diagnostic biopsies stratified by immune response, where patients with an early response (red) exhibited significantly lower TEX activity compared with those with a late (blue) or no response (black) (p = 0.026). **(F)** Kaplan–Meier analysis of PCSM in the UV1 cohort stratified by the median GSVA score for the TEX signature is shown, with the two-sided log-rank p-value above the curves (p = 0.02).

The lack of an association between AR activity and clinical outcome in the IMRT cohort suggests that AR status alone is insufficient to predict therapeutic response, and that combined assessment of AR signaling and immune status may be required to inform the use of ADT in combination with UV1 vaccination. This is supported by previous reports showing AR activity as a strong predictor of durable response to AR pathway inhibitors (10,27).

Given the prominent role of immune-related classifiers such as association between TEX and outcomes in the UV1 cohort, to validate the contribution of T cell exhaustion, we next examined the association between clinical outcome and 142 gene signatures depicting T cell exhaustion, described in published meta-analysis (28). In line with our earlier findings, 18.3% (26 out 142 signatures) of the signatures analyzed revealed an association with poor prognosis exclusively in the UV1 cohort against PCSM (**Figure 3C)** and PFS (**Supplementary figure 4C),** and not in the IMRT cohort (**Figure 3D** and **Supplementary figure 4D**).

To corroborate whether baseline T-cell dysfunction influences clinical outcome following UV1 hTERT vaccination, we assessed the TEX gene expression signature scores in pre-treatment tumor samples from the UV1 cohort. TEX signature scores varied significantly across immune response groups (**Figure 3E**). Patients who subsequently exhibited an early clinical response had markedly lower TEX signature activity at baseline compared with patients with late or no response (*p* = 0.03), indicating that reduced T-cell exhaustion prior to treatment is associated with improved responsiveness to vaccination. Consistent with this observation, low TEX score at baseline was significantly associated to longer time to PCSM (*p* = 0.02; **Figure 2F**) and PFS (*p* = 0.04; **Supplementary Figure 5B**) compared with patients exhibiting high TEX score. These associations were specific to the UV1 cohort and were not observed in the IMRT cohort (**Supplementary Figure 5A and C**).

Collectively, these findings demonstrate that the pre-treatment functional state of intratumoral T cells, specifically the degree of exhaustion, is a major determinant of both vaccine-induced immune responses and long-term clinical outcomes.

### CD8⁺ T-cell abundance is associated with clinical outcome following UV1 vaccination

To characterize the immune cells composition associated with response to UV1 hTERT vaccination, we performed gene expression deconvolution of tumor tissue using EPIC (15). This analysis revealed a significant increase in the fraction of CD8⁺ T cells in post UV1 treatment biopsies compared with matched pre-treatment diagnostic biopsies (p = 0.004; **Figure 4A**), indicating enhanced intraprostatic T cell infiltration following UV1 treatment initiation. Supporting these results, the CD4 inferred aboundance exhibited a positive correlation with TEX score, while the CD8 score was inversely correlated (**Figure 4B**). Consistently, higher baseline CD8⁺ T-cell abundance, was strongly associated with longer time to progression (p = 0.003; **Figure 4C**) and PC-specific death (p = 0.0005; **Figure 4D**).

**Figure 4.**
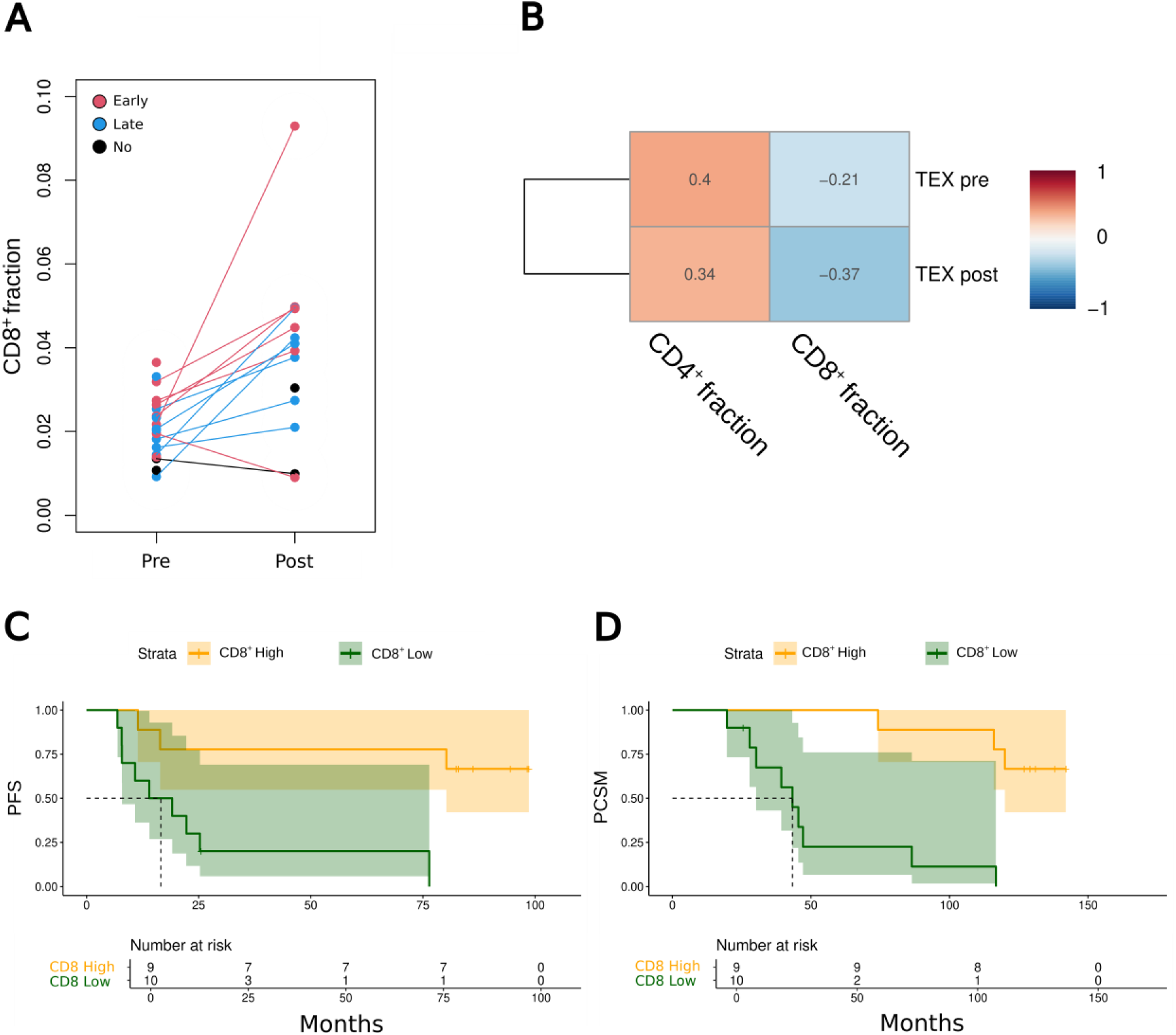
CD8⁺ T-cell abundance is associated with clinical outcome following UV1 vaccination. **(A)** CD8⁺ T-cell fraction inferred from gene expression deconvolution inferred with the tool EPIC (See Methods) in paired pre-and post-UV1 treatment samples. Post-treatment samples exhibited a significantly higher T-cell fraction compared with pre-treatment samples (*p* = 0.004). **(B)** Correlation analysis of exhausted T-cell (TEX) signature activity with EPIC-inferred T-cell fractions before and after vaccination in the UV1 cohort. **(C)** Kaplan–Meier analysis of progression-free survival (PFS) for UV1 cohort stratified into two groups based on inferred CD8⁺ T-cell fraction inferred from gene expression deconvolution. The two-sided log-rank p-value is shown above the curves (p = 0.003). **(D)** Kaplan–Meier analysis of prostate cancer-specific mortality (PCSM) for UV1 cohort stratified into two groups based on inferred CD8⁺ T-cell fraction inferred from gene expression deconvolution. The two-sided log-rank p-value is shown.

### Distinct T cell repertoire dynamics differentiate early and late immune responders to UV1 vaccination

Given that the UV1 peptides were developed to elicit T-cell response, we inferred prostate infiltration of T-cells post vaccination. As higher CD8+ presence in diagnostic biopsies was associated with longer PFS and PCSM, we sought to investigate whether TCR-repertoire was skewed toward specific sequences, reflecting potential tumor killing activity due to the vaccination. For these reasons, we considered two parameters: TCR clonal diversity and evenness in pre-and post-treatment biopsies (**Figure 5**).

**Figure 5.**
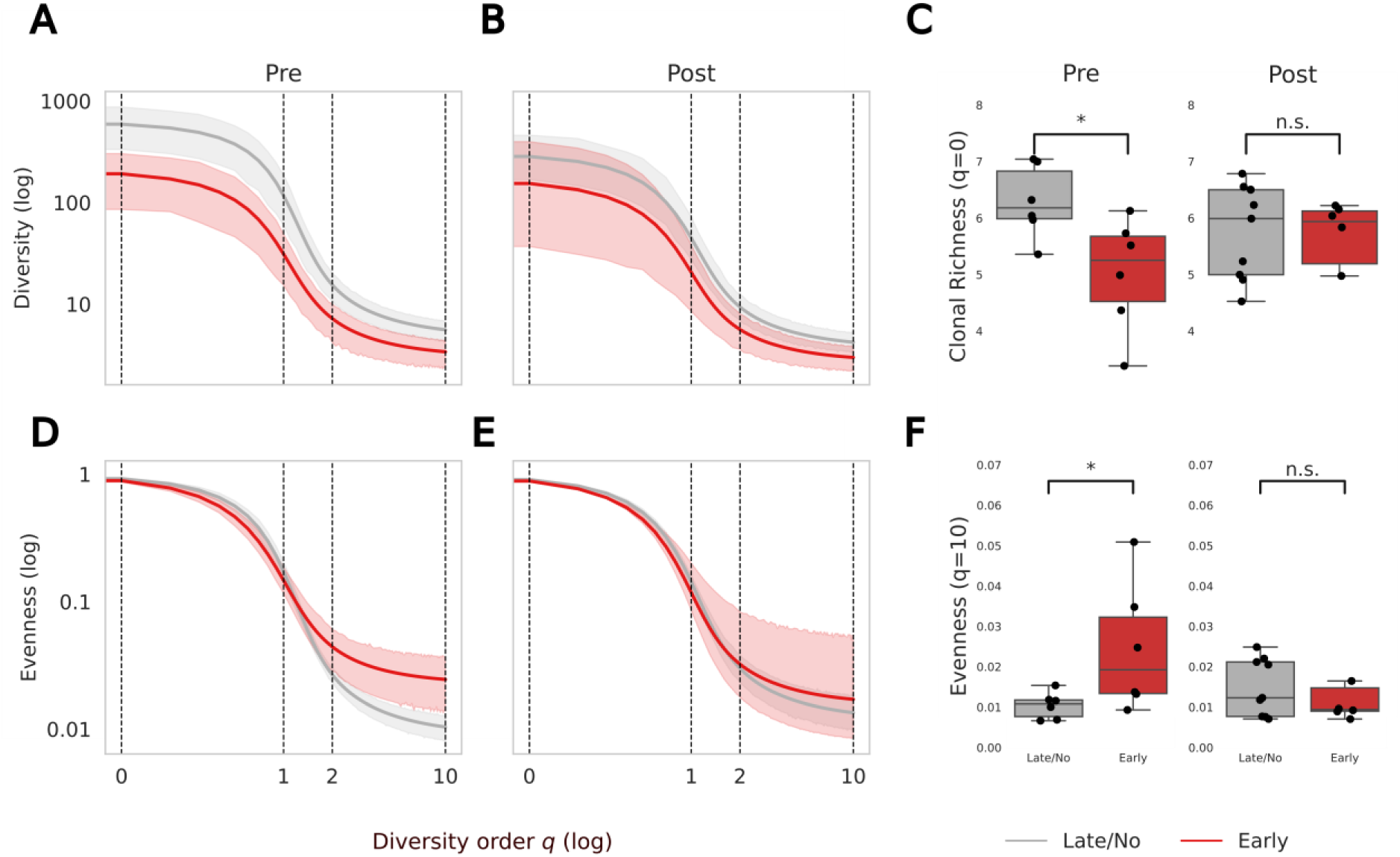
TCR clonal diversity and evenness are determinants of immune responses to UV1 vaccine. (A-B) Hill diversity profiles in pre-UV1 treatment diagnostic biopsies (A) and post-UV1 treatment biopsies **(B)** across a range of diversity orders (*q*). At diversity order 0, diversity equals the total number of unique TCR motifs (clones) detected in the sample while at higher diversity orders, rare clones contribute less, accounting for differences in sampling or sequencing depth between sample types. **(C)** Boxplots of total TCR motifs (clonal richness) comparing pre-UV1 treatment and post-UV1 treatment samples. Asterisks indicate Welch’s t-test *p* < 0.05. **(D-E)** Hill evenness profiles in pre UV1 treatment diagnostic biopsies **(D)** and post UV1 treatment biopsies **(E)** calculated by dividing the diversity profile by its value at Hill diversity order q = 0 (total number of clones in the repertoire). Curves that drop further from baseline at higher *q* values indicate more skewed, or uneven, distributions. Both evenness (y-axis) and diversity order (x-axis) are log-transformed. **(F)** Boxplots of evenness at diversity order q = 10, where lower values indicate more skewed TCR repertoires while higher values indicate more evenly distributed T cells. Asterisks indicate Welch’s t-test *p* < 0.05.

We assumed that a clonally expanded TCR repertoire was characterized by a highly uneven clonal frequency distribution, in which a small subset of clones accounts for the majority of TCR sequences detected in the sample. We used Hill diversity and evenness profiles (17) to assess the skewness of the clonal frequency distributions, where the diversity of the repertoire was calculated across a range of diversity orders, *q* (See Methods). At higher values of q, diversity calculations give less weight to rare clones, allowing assessment of whether observed differences in diversity between samples persist despite variations in sampling or sequencing depth (see Methods). Evenness, meanwhile, reflects how uniformly TCR sequences are distributed across clones—and, in our analysis, across the motifs derived by the TCR sequences (see Methods). High evenness indicates an unexpanded repertoire, whereas low evenness suggests clonal expansion.

In tumor tissue samples collected prior to vaccination, early immune responders exhibited higher TCR motif diversity across all diversity orders compared to late/no responders (**Figure 5A-C**). At diversity order q = 0 (species richness), late/no responders had a median motif diversity of 488.2 (range: 212.3–1141.6), compared to 197.8 (range: 29.5–458.7) (p = 0.03) in early responders, suggesting that, prior to vaccination, early responder tissues contained fewer unique clonotypes infiltrating the tumor. This difference was no longer significant in post-vaccination samples (**Figure 5B-C**), indicating that determinants of immune response were present prior to vaccination. Early immune responders showed higher evenness at higher diversity orders (q > 2) in pre-vaccination samples compared to late/no responders (**Figure 5D-F**), a pattern that disappeared in post-vaccination samples (**Figure 5E-F**).

These results suggest that tissue infiltrating T-cell clones in early immune responders were uniformly distributed in terms of T-cell counts yet had not shifted toward an exhausted phenotype prior to the UV1 vaccination.

### Pre-vaccination biopsies of early immune responders are enriched for a distinct set of TCR sequence motifs

To explore whether specific TCR sequences of infiltrating T-cells prior to the UV1 vaccination could explain differences observed in the immune response, we next analyzed TCR motifs to determine if they could stratify early and late or non-immune responders. Clustering the TCR motifs in pre-UV1 treatment biopsy samples revealed two main clusters which clearly separated the early versus late/no immune responders (**Figure 6A**). This clear separation of patients based on TCR motifs suggests that pre-existing differences in TCR repertoire underlie the ability of patients to mount an effective response to UV1 vaccination. No such separation was observed in post UV1 vaccination tissue samples based on TCR motifs **(Supplementary Figure 6).**

**Figure 6.**
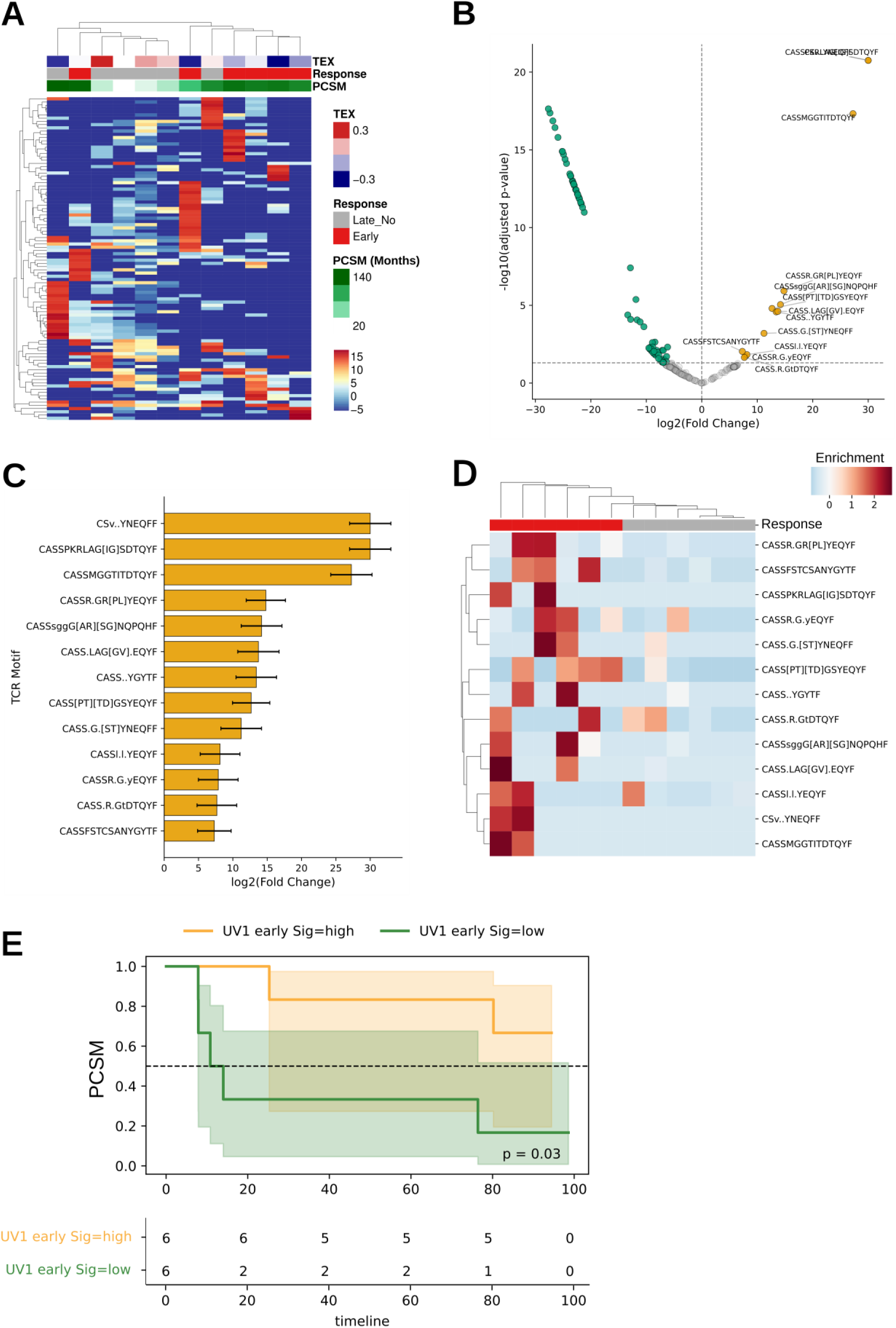
Distinct TCR sequence motifs enriched in pre-vaccination repertoires are associated with subsequent response to UV1 vaccination. **(A)** Cluster heatmap of core needle biopsies taken prior to UV1 vaccination. Values represent clusTCR-derived sequence motifs, hierarchically clustered using complete linkage and Euclidean distance, based on variance-stabilized counts. Prior to clustering, motifs present in fewer than three samples with at least one count were filtered out, and the top 100 motifs by variance were selected. The heatmap columns are annotated with Response, Prostate Cancer Specific Mortality (PCSM). **(B)** Volcano plot of DESeq2 differential expression between early responders and late/non-responders to UV1 vaccination. The x-axis shows log₂ fold changes, with positive values indicating higher expression in early responders. The y-axis shows the –log₁₀ FDR for each motif. Motifs significantly enriched in early responders are highlighted in yellow, with their sequences labeled on the plot. Depleted motifs are highlighted in green. **(C)** Bar plots of log₂ fold changes for motifs enriched in early responders. The x-axis shows the log₂ fold change, and error bars represent the standard error of the fold change. **(D)** Cluster heatmap of motifs enriched in early responders. Values were converted to counts per million (CPM), log-transformed, and then z-score normalized across samples for each motif. **(E)** Kaplan–Meier analysis of PCSM comparing samples stratified into high versus low abundance of early UV1 immune responder TCR signature motifs.

Next, we performed a differential motif abundance analysis of pre-vaccination TCR motifs and we identified 13 motifs significantly enriched in early immune responders (FDR < 0.05) compared to late/no immune responders (**Figure 6B-C**). While these motifs were not uniformly highly expressed across all early responders, each motif was enriched in at least two early responder samples (**Figure 6D**), highlighting both shared and patient-specific features of the pre-existing TCR repertoire. Overall, these findings support the concept that baseline intratumoral TCR architecture, including motif composition and T-cell exhaustion status, may contribute to the capacity of a cancer vaccine-induced immune response, and that specific clonotypes are preferentially associated with favorable patient outcomes. Kaplan–Meier curves confirmed that patients with high early-signature TCR motif abundance had improved PCSM compared with low-abundance patients (p = 0.03; **Figure 6E**).

To understand whether tissue-derived TCR repertoires within the systemic immune response reflected similar expansions observed in the prostate tissue, we also analyzed the TCR repertoire of T cells isolated from PBMCs of the same patients, collected at several intervals pre and post UV1 vaccination. Pairwise sample repertoire TCR motif overlap analysis using the Morisita–Horn Index (MOI) (see Methods), which intrinsically accounts for motif frequencies within each compared repertoire, revealed, as expected, minimal overlaps between TCR repertoires from different patients (**Supplementary Figure 7A**). The overall overlap of TCR repertoires between biopsies and PBMCs was generally low (**Supplementary Figure 7A**), suggesting caution when using PBMCs to interpret the tumor immune response.

In contrast, within-patient comparisons showed marked differences in clonal overlap depending on whether samples originated from PBMCs or prostate tissue (**Supplementary Figure 7B**). TCR repertoires from PBMCs sampled longitudinally—from baseline (prior to UV1 vaccination) through follow-up—showed consistently higher MOI values (median MOI=0.76, two-sided Mann–Whitney U test: *U* = 162, *p* = 9.34 × 10⁻⁵) (**Supplementary Figure 7C**) than paired tissue samples (**Supplementary Figure 7D**) median MOI=0.21). Although patient-specific, the consistent within-patient overlap between TCRs from prostate tissue and PBMCs motivated a longitudinal analysis to identify T cell clones that expanded following vaccination, and to the assessment of their persistence over time in the PBMCs.

Six thousand one hundred ninety-six out of 529,299 (1.2%) TCR clonal motifs expanded in PBMCs from baseline to the first phlebotomy, defined by a log₂ fold change >2 and persistence in over 50% of subsequent samples (**Supplementary Figure 8A**). To assess whether these clones were vaccine-associated, we performed a sequence convergence analysis using Levenshtein distance (LD), constructing LD-based graphs from the underlying TCR sequences. Sampling 5000 expanded or randomly selected TCRs 100 times each, all graphs contained ∼5000 nodes, but the number of edges between similar TCR sequences differed markedly: expanded samples had a median of 448 edges, compared to 168.5 in random samples (two-sided Mann–Whitney U test: U = 10,000, p = 2.53 × 10⁻³⁴) (**Supplementary Figure 8B**). Consistent with antigen-driven responses producing convergent TCRs, expanded clones showed significantly greater convergence than expected by chance.

Together, this observed expansion of TCR sequences, with sustained persistence and increased sequence motif convergence, indicate that UV1 vaccination induced antigen-driven T cell clonal responses in peripheral blood.

### UV1 vaccine–associated T-cell receptor motifs are detected in prostate tumor regions of non-vaccinated patients, yet are absent in ADT resistant tumors

To investigate the intratumoral localization of TCR motifs associated with vaccine response, we employed spatial VDJ (21) to map TCR repertoires with spatial references to the regions within the tissue that encoded specific CDR3 regions. This allowed defining spatial distribution of TCR sequences in prostate tissue sections using long-read spatial VDJ sequencing (See Methods). Analyses were performed on specimens from non-UV1–treated patients (**Figure 7A**) comprising one PC patient treated with neoadjuvant ADT (NEADT), one treatment-naïve prostate cancer (TRNA) patient, one patient undergoing prostatectomy for benign prostatic hyperplasia (BPH), and one patient undergoing transurethral resection of the prostate for relapsing castration-resistant prostate cancer (CRPC). For these specimens, spatial transcriptomics data and single-cell–derived annotations (SCM) of major tissue compartments spot level—including tumor, immune, basal, luminal, club-like, fibroblast, endothelial, and muscle regions—were available (**Figure 7B**) (22).

**Figure 7.**
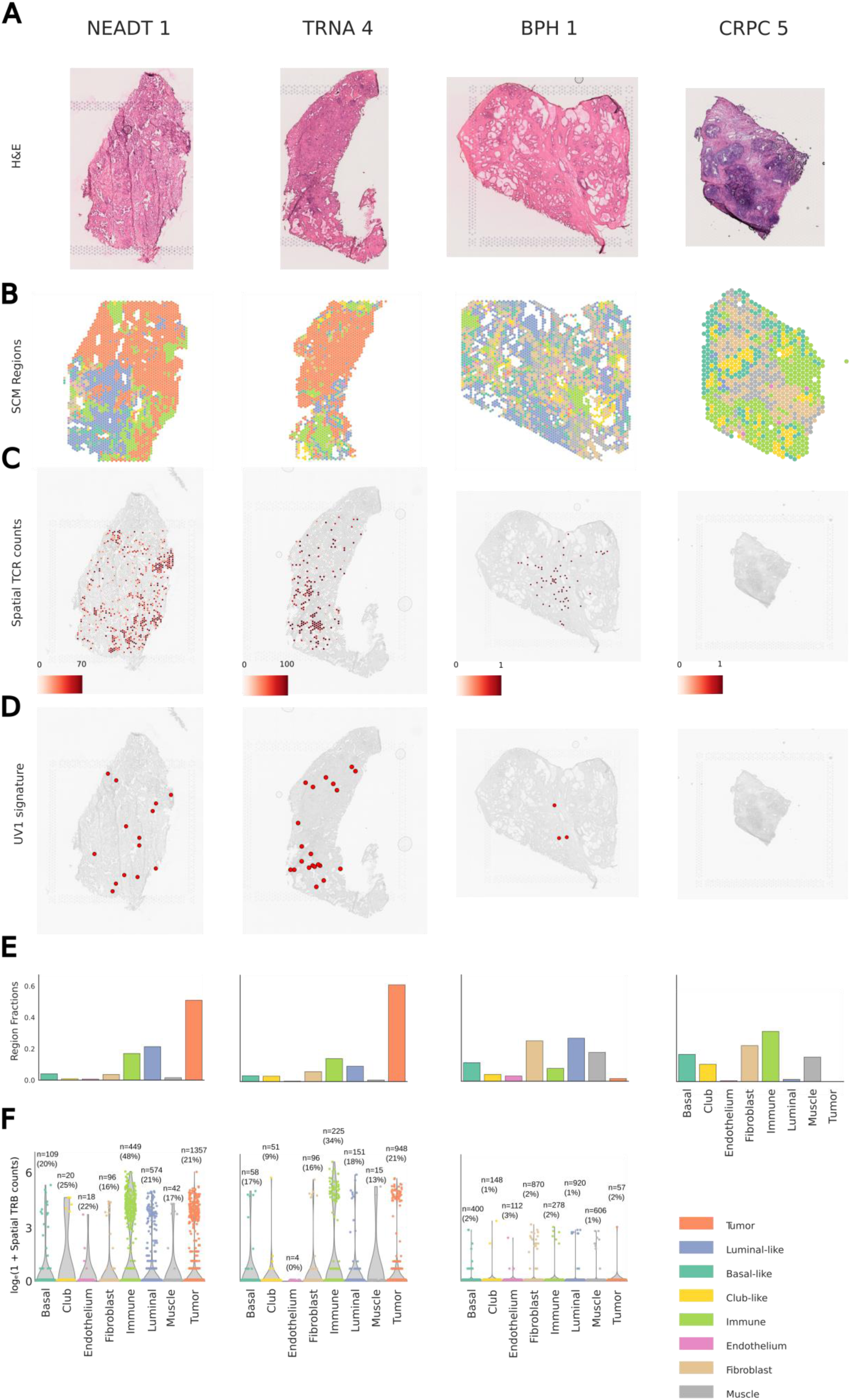
TCR sequence motifs associated with immune response to UV1 vaccination are detectable in prostate cancerous tissue. (**A**) H&E-stained tissue sections from neoadjuvant-treated prostate cancer (NEADT), treatment-naïve prostate cancer (TRNA), treatment-naïve benign prostatic hyperplasia (BPH), and castration-resistant prostate cancer (CRPC). (**B**) Single-cell mapping–derived region-(SCM) annotations overlaid on each spatial transcriptomics spot, with regions annotated as tumor, basal-like, immune, fibroblast, luminal-like, club-like, endothelium, or muscle according to Kiviaho et al. 2024 (22)(22) (**C**) Overlaid absolute counts of PacBio long sequencing reads mapping to CDR3 loci containing both V(D)J recombination sequences and spatial barcodes. The colormap represents the raw number of spatially barcoded CDR3-mapped reads containing the spatial coordinate at that location. (**D**) Red dots indicating spatial mapping of T cell receptors overlapping the differentially enriched TCR motifs that distinguish pre-diagnostic biopsies of the UV1 trial patients. A spatial coordinate was highlighted if at least one read contained an aligned TCR sequence matching a UV1 signature motif (**E**) Fraction of spatial spots assigned to each SCM region (basal-like, club-like, endothelium, fibroblast, immune, luminal-like, muscle, and tumor) for each tissue section. (**F**) Overlap of spatial VDJ TCR counts per spot, stratified by SCM region. Counts are log-transformed with an added pseudo count. N represents the total number of spatial spots within the annotated SCM region, while the percentage indicates the fraction of those spots containing at least one spatial TCR read.

Spatial VDJ sequencing revealed marked differences in TCR abundance across samples (**Figure 7C**). Strikingly, TCR sequence motifs associated with early immune response to UV1 vaccine were detectable among the repertoires (**Figure 7D**). Together with other TCRs sequences, these motifs closely tracked tumor content in the specimens, indicating that UV1 treatment-naïve patients develop clonal expansions that could potentially mark their response to the vaccination (**Figure 7D**). SCM annotations for the profiled specimens revealed that tumor regions constituted 51.0% and 61.1% of spatial spots in the NEADT and TRNA samples, respectively, whereas only 1.7% of spots were classified as tumor in the BPH sample, and no tumor regions were found in this particular CRPC sample (**Figure 7E**).

Nevertheless, cancerous spots annotated as “club-like” were present in all samples and were significantly more abundant in the CRPC specimens. Interestingly, and corroborating our previous finding that club-like cells define a cancerous phenotype, associated with myeloid-mediated immunosuppression, treatment resistance, and found also in metastatic specimens (22), the CRPC specimens had massive infiltration of immune cells, but most likely neutrophils (31.7% of the spots) as previously reported (22) but no T or B cells, as opposed to 17.0% of spots in NEADT, 14.5% in TRNA, 8.1% in BPH which, although annotated generally as “immune” had substantial infiltration of T and B cells.

TCR sequencing reads were concordant with previously inferred region annotations, with immune-annotated spots generally showing higher or comparable TCR counts to tumor-annotated spots in the NEADT and TRNA samples, while very few or no TCR reads were detected in BPH and CRPC samples (**Figure 7F**). NEADT and TRNA sections had detectable T cell infiltration with motif-aligned TCRs spatially co-localizing with regions of high T cell density (**Figures 7B and D)**. In the BPH samples, the scarcity of TCR reads mirrored the limited tumor content indicated by the SCM annotations.

Given the vast diversity of TCR sequences and the low degree of sequence overlap between patients (**Supplementary Figure 7A**), the reproducible spatial detection of motif-aligned TCRs across samples highlights their potential as robust indicators of tumor immune contexture.

Together, these findings show that UV1 vaccine–responsive T-cell receptor sequence motifs are detected in UV1-naïve prostate cancerous regions pre and post ADT, and demonstrate that TCR motif-informed spatial mapping can provide critical insight into the tissue-level determinants of cancer vaccine responsiveness.

## Discussion

The single-arm phase I/II clinical trial evaluating UV1 vaccination (NCT01784913) demonstrated safety in patients with de novo mCSPC (7). However, UV1 vaccination yielded disappointing results in several clinical trials, failing to demonstrate clinical benefit in metastatic or recurrent head and neck squamous cell carcinoma and melanoma when combined with pembrolizumab (29) (NCT05075122), as well as unresectable metastatic melanoma and malignant pleural mesothelioma in combination with immune checkpoint inhibitors (ICI) (29). Here we show that low volume mCSPC patients experiencing early immune response to the vaccine had an exceptionally favorable 10-year survival rate, comparable to patients diagnosed with non-metastatic disease.

In mCSPC, the primary unmet clinical challenge is the eradication of minimal residual disease following otherwise relatively effective standard-of-care treatments. Therefore, differences in low versus high volume metastatic patients can be the basis for the remarkable benefit observed in this PC trial compared to the other trials.

ICIs have been used in metastatic CRPC patients with disappointing results (30,31) due to intrinsic features of the tumor immune microenvironment (32). However, in the NEPTUNES trial, a selection of mCRPC patients with favorable immune profiles was associated with improved responses to ICI (33).

In this study, we show that although patients treated with UV1 were not selected based on pre-existing immunological or molecular biomarkers, they exhibited molecular differences distinguishable in outcomes, suggesting that a subset of mCSPC patients can be selected for UV1 vaccination, and plausibly even for ICI, or other types of autovaccinations (34) based on pre-existing tumor-immune characteristics. Specifically, analysis of immune-related features associated with clinical response and survival following UV1 vaccination indicates that therapeutic efficacy depends on the presence of non-exhausted resident intratumoral T cells (34). These observations point to the possibility that AR inhibition may contribute to T cell exhaustion, potentially by altering AR signaling activity within immune cells (10,14,35). The pattern of TCR expansions in the prostate biopsies before and after vaccination and from the PBMCs suggests pronounced immunoediting. Notably, the pre-vaccination TCR repertoire separation between tissue samples from early and late responders indicate that TCR repertoire profiling provides higher-resolution insight into intratumoral immune dynamics than PBMC-derived repertoires. In PBMCs, the high clonal overlap across time points in PBMCs within the same patient, together with low overlap between patients, is expected, due to long-term memory from prior exposures to vaccines, viruses, and other pathogens, as well as the large diversity of circulating clones, most of which are unlikely to participate directly in the UV1 vaccine response.

Together, these findings indicate that to understand response to cancer vaccine, it is paramount to study tumor immunoediting through TCR repertoires analyses. Therapeutic combinations involving immunotherapies have been shown to induce distinct T-cell receptor (TCR) repertoire responses across multiple malignancies (36), and TCR repertoire–based clonality has demonstrated prognostic value in diverse clinical settings (11). Although identification of TCR sequences specific for hTERT-derived antigens remains technically challenging with existing approaches, our study revealed meaningful differences in clonal frequency distributions that distinguished early responders from late or non-responders prior to vaccination.

These differences included specific TCR sequence motifs associated with response, which were also detectable in a tissue-spatial context in an independent cohort of PC patients with no prior exposure to the vaccine. Our findings indicate that a favorable early immune response to the vaccine is predetermined by pre-existing features of the patients TCR repertoire. Moreover, increased sequence convergence among persistently expanded clones in post-vaccination peripheral blood supports antigen-driven clonal expansion, with expanded clonotypes detectable in both prostate tissue and circulation. The detection of these repertoire features in patients not pre-selected based on immunological or molecular characteristics suggests the presence of endogenous immune responses to hTERT-derived antigens within the prostate and warrants further investigation.

Spatial VDJ profiling also revealed tissue-level organization of such vaccine response-associated TCR repertoires, indicating that overall tissue composition strongly influences the extent of T cell infiltration, with the presence of tumor tissue being critical for some immuno-engagement.

The UV1 vaccine enhances the response to endogenous antigens, and it is effective in limiting metastatic seeding, consistent with its design to engage CD4+ T helper cell-mediated immune functions, which renders it a particularly attractive strategy in mCSPC because of minimal toxicity compared to androgen receptor pathway inhibitors, and because of a durable response. Even if, based on our analysis, we could not clearly distinguish CD4+ T cell activation from the overall T cell response, supporting this observation, ADT has been associated with increased T helper and fewer CD8+ T cells infiltration in the prostate (37). Furthermore, a higher circulating tumor cells burden was associated with better survival rate in the same trial (7) and corroborates our data in suggesting that patients with a primed immune system derive greater benefit from vaccination.

Taken together, our findings show that UV1 vaccination amplifies pre-existing antitumor immune activity rather than initiating de novo immunity, exerting its greatest benefit in the setting of low metastatic volume and minimal residual disease.

## Funding

AU is K. Albin Johansson Cancer Research Fellow and Tampere Institute of Advanced Study Research Fellow. AU wishes to thank Finnish Cancer Institute, the Research Council of Finland project no. 349314; Cancer Foundation Finland; Norwegian Cancer Society project no. 198016-2018 & project no. 273672 –2023 and Radiumhospitalets Legater. EH wishes to thank Norwegian Cancer Society project no. 273672 –2023; TC wishes to thank EDUFI and Pirha VTR program, KS wishes to thank the Tampere Institute of Advanced Study, SE wishes to thank the Cancer Foundation Finland and the Research Council of Finland project no. 349314. MN wishes to thank the Academy of Finland Center of Excellence program (project no. 312043), the Sigrid Jusélius Foundation, and the Cancer Foundation Finland. WL wishes to thank the Radiumhospitalets Legater, and the Raagholtstiftelsen.

## Data Availability

All data produced in the present study are available upon reasonable request to the authors

## Acknowledgments

We are deeply grateful to the patients and their families for their participation in these clinical trials. We thank the Tampere Genomics Facility, the DNA Sequencing and Genomics Laboratory (BIDGEN) at the Institute of Biotechnology in Helsinki, and the Oslo NorSeq sequencing Facility at the Oslo University Hospital for the support for sequencing the clinical samples. We are grateful to Cegat GmBh, and Enpicom for sequencing and data processing services with the TCR sequencing. We acknowledge Covaris LTD for the support on the extraction of nucleic acids from FFPE samples.

## Data and code Availability

All data supporting the findings of this study are available in the paper and its Supplementary Information section. Given the sensitivity of the data generated, raw data are available upon reasonable request. The gene expression count matrix is available as **Supplementary File 1.** Any additional information required to reanalyze the data reported in this paper is available from the lead contact upon request. All analysis code used in this study is available at GitHub (https://github.com/eirikhoye/uv1-prostate-transcriptome-tcr).

## Contributions

AU conceived and designed the study, provided resources, and supervised the project. EH performed experiments and data analyses. EH, RN, and AU drafted the manuscript and analyzed and interpreted the data. KS and SE generated spatial V(D)J sequencing data, and TC contributed to its generation. NE and JES generated the RNA-seq and TCR-seq data. SH, AK, and FB performed bioinformatics analyses. TV, TT, TJM, WL, MN, and AM provided resources and clinical material. WL served as principal investigator of clinical trials and managed the clinical data. All authors revised the manuscript and approved the final version.

## Notes

### Competing Interest Statement

MW is an employee at Ultimovacs and EBE has been employed by Ultimovacs, the company that developed the UV1 vaccine. Teemu J Murtola has received consultant fees from Astellas, Johnson & Johnson, Amgen, Pfizer and Astrazeneca; lecture fees from Bayer, Roche, Pfizer, Ipsen, Astellas, Amgen, and Johnson & Johnson; and research funding from Bayer, Pfizer and Johnson & Johnson. The remaining authors declare no competing interests.

### Clinical Trial

NCT01784913

### Author Declarations

The Regional Committee for Medical Research Ethics-Southeast Norway gave ethical approval of this work (S-2019/1790).

